# Efficacy of Radiotherapy in Osteoarthritis: Survey of Orthopaedic Surgeons, Hand Surgeons, General Practitioners, and Radiotherapists

**DOI:** 10.1101/2025.06.15.25328966

**Authors:** Helmut H. Huberti

## Abstract

**Background:** Low-dose radiotherapy (LD-RT) has been reported to alleviate symptoms in osteoarthritis (OA) by modulating soft tissue inflammation. While supported by experimental and guideline-based evidence, its clinical effectiveness in everyday practice remains debated, particularly in Germany.

**Methods:** A nationwide postal survey was conducted among 6,943 practicing physicians in Germany, including orthopaedic surgeons, hand surgeons, general practitioners, and radiotherapists. A standardized three-question questionnaire assessed estimated annual LD-RT usage, rates of symptom resolution, and the need for a second treatment series. Responses were returned via fax and analyzed descriptively.

**Results:** A total of 618 physicians (response rate: 8.9%) reported treating 37,754 OA patients annually with LD-RT, averaging 61 patients per physician (median: 20). Across all respondents, the mean reported rate of long-term symptom relief was 45%, with a second LD-RT series administered in 47% of cases. Reported effectiveness varied by specialty: general practitioners (59%), radiotherapists (58%), orthopaedic surgeons (44%), and hand surgeons (31%).

**Conclusions:** This physician-reported survey suggests that LD-RT is perceived as a moderately effective intervention for osteoarthritis, typically used as a last-resort therapy. Its reported effectiveness in nearly half of treated patients supports the rationale for broader clinical evaluation, and potentially earlier use in the treatment course. Further prospective trials are needed to validate these observational findings.

## INTRODUCTION

Over several years, the German Osteoarthritis Society (DAH, Deutsche Arthrose-Hilfe e.V.) has informed and advised more than 2.5 million osteoarthritis patients in four countries about all aspects of osteoarthritis and polyosteoarthritis. In its full manifestation, e.g., the latter causes pain in all fingers of both hands, leads to major limitations in daily life, alters appearance, and results in enormous psychological distress for many—mostly female—patients. Standard local medical interventions can alleviate symptoms but usually fail to stop disease progression.

Repeatedly, patients have reported positive experiences with radiotherapy. It is scientifically known that radiotherapy with low-dose X-rays (“Low-Dose Radiotherapy,” or “LD-RT”) can modulate, reduce, or completely eliminate soft tissue inflammation through molecular and radiobiological mechanisms. This is supported by numerous experimental studies and the evidence-based guidelines issued by the German Radiation Oncology Society (DEGRO) (1). However, in medical practice the clinical effectiveness of LD-RT in osteoarthritis still remains a subject of debate.

Also, a new literature review on Heberden’s osteoarthritis (2) revealed inconsistent findings that does not allow a definitive answer regarding the clinical efficacy of LD-RT in finger osteoarthritis—particularly since the studies were too small. However, the absence of evidence does not equate to proof of no effect. Could LD-RT in fact be a particularly effective treatment for osteoarthritis, perhaps even an outstanding method of last resort? Could physician-based DROM surveys (Doctor Reported Outcome Measures), given large sample sizes and specific focus, offer clear answers?

## OBJECTIVES

The objectives of this study were to conduct comprehensive, nationwide surveys among nearly all practicing orthopaedic surgeons, hand surgeons, radiotherapists, and a significant number of general practitioners in Germany about their own usage frequency and observations regarding the effectiveness of LD radiotherapy in osteoarthritis.

## METHODS & MATERIALS

We surveyed a total of 6,943 practicing physicians about their experience with radiotherapy in osteoarthritis. Specifically, these included 4,645 orthopaedic surgeons, 1,068 hand surgeons, 1,000 general practitioners, and 230 radiotherapists. Addresses of the orthopaedic surgeons and general practitioners were rented from the company SciTrace GmbH, Tübingen. The contact information for hand surgeons and radiotherapists was collected and copied from internet sources.

All physicians received a one-page questionnaire containing three questions: For orthopaedic surgeons and for hand surgeons:

1. For how many of your osteoarthritis patients is radiotherapy performed each year? *N = per year (estimated)*
2. In approximately what percentage of cases does this completely and permanently relieve pain and soft tissue swelling? *Approximately % of cases (estimated)*
3. In approximately what percentage of cases is a second RT series conducted? *Approximately % of cases (estimated)*

For general practitioners and radiotherapists, question 1 used the phrase “your patients with finger joint osteoarthritis” instead of “your osteoarthritis patients.” Otherwise, the questions were identical.

Questionnaires were sent in print by postal mail. Responses were requested by fax. The data were entered manually into Excel spreadsheets, including the date received, address ID, and the three responses. Evaluation was conducted using Excel functions for calculating means, frequency distribution histograms, and medians.

## RESULTS

### Overall Overview

Of the 6,943 physicians surveyed, 618 (8.9%) responded. Each year, they treat a total of 37,754 osteoarthritis patients with radiotherapy—an average of 61 patients per physician (range: 0–1000), median 20. Thus, the responding physicians apply radiotherapy to approximately one patient per week.

In nearly half of the patients, low-dose radiotherapy leads to long-term relief of symptoms. The average success rate was 45% (range 0–100%), median 50%. A second LD-RT series is administered in 46.8% of cases on average (median: 50%).

### Results by Physician Group

- **409 orthopaedic surgeons** (8.8% of those surveyed) estimated that they treat a total of 22,173 osteoarthritis patients with LD-RT per year. The average per physician was 54 (0–1000), median 20. Long-term relief was achieved in 44.4% of patients (median 50%). A second LD-RT series was performed in 48% of cases (median 50%).
- **102 hand surgeons** (9.6%) reported 3,910 patients per year, average 38 (0–700), median 10. Long-term relief was achieved in 30.7% of cases (0–80%), median 25%. A second LD-RT series was used in 39% of cases (0–100%), median 30%.
- **35 general practitioners** (3.5%) reported 1,129 patients, average 32 (0–1000), median 2. Long-term relief was achieved in 58.9% of cases (0–100%), median 55%. A second LD-RT series was performed in 32% of cases (0–100%), median 30%.
- **72 radiotherapists** (31.3%) reported 10,543 patients, average 146 (10–800), median 80. Long-term relief was achieved in 57.5% of cases (0–90%), median 60%. A second LD-RT series was used in 52% of cases (0–100%), median 50%.

### Table

| **Group** | **Contacted** | **Respondents** | **Patients /**<br>**year** | **Effectiveness**<br>**(mean)** | **2nd Series**<br>**(mean)** |
| --- | --- | --- | --- | --- | --- |
| All | 6,943 | 618 (10.3%) | 37,754 | 45% | 47% |
| Orthopaedic | 4,645 | 409 (8.8%) | 22,173 | 44% | 50% |
| Hand Surgeons | 1,068 | 102 (9.6%) | 3,910 | 31% | 39% |
| General Pract. | 1,000 | 35 (3.5%) | 1,129 | 59% | 32% |
| Radiotherapists | 230 | 72 (31.3%) | 10,543 | 58% | 52% |

## DISCUSSION

This study included only three questions aimed at the four physician groups who treat the majority of osteoarthritis patients in Germany. Question 1 varied slightly among the groups. For orthopaedic surgeons, it addressed all osteoarthritis patients regardless of the affected joints. For hand surgeons, the question naturally focused on hand osteoarthritis. For general practitioners and radiotherapists, the question explicitly referred to finger joint osteoarthritis. These differences must be considered when comparing the results, but they are of minor importance for the core findings of this study.

In response to question 2, 618 German physicians stated in writing that radiotherapy (LD-RT) can completely and permanently eliminate pain and soft tissue swelling in osteoarthritis. They also noted that this effect was observed in about every second or third treated patient. This applied both to general osteoarthritis (orthopaedic surgeons) and to hand and finger osteoarthritis (hand surgeons, general practitioners, radiotherapists).

Surveys as a research method typically show high interindividual variability, which is also seen in the present responses—ranging from 0% (never effective) to 100% (effective in all patients). Nevertheless, mean and median values provide meaningful insights. The key questions—”Is LD-RT clinically effective?” and “How likely is it to work?”—can be reliably answered. These two pieces of information are essential for patients. Those suffering from chronic daily pain and limitations are likely to accept the logistical effort (six sessions in two weeks, possibly a second series) for a 50% chance of success.

Whether the true probability is 50%, 43%, or only 30% is of secondary importance to some extent and also depends on many yet undefined factors. As seen in the median values, most physicians treat only a few patients per year with LD-RT. A smaller number, however, treat up to 1,000 patients annually, corresponding to 3–4 patients per day. That’s a notably high number, even for a large practice. The variation in usage reflects not only experience with the method but also influences further indications. A physician who uses RT only once a year and sees no effect may feel validated in their caution, and vice versa.

The fear of serious side effects from X-ray therapy is historically rooted. When observing the age limit of 50 years (DAH recommends 60), this caution is no longer justified (1).

The more often this is acknowledged, the more the role of LD-RT as a last resort may shift. The earlier LD-RT is used, the more full-blown osteoarthritis cases might be avoided—and the more favorable the efficacy statistics may become. LD-RT primarily targets soft tissue inflammation and probably has significantly less impact on bone-related pain and limitations in advanced stages. The results of question 3—showing that on average, a second irradiation series was needed in 47% of patients—may also support this assumption.

## CONCLUSION

Based on the data presented, we draw the following conclusions:

1. LD-RT is currently a last-resort therapy for osteoarthritis, used only when all other methods have failed—typically in later stages of the disease.
2. The effect of LD-RT does not occur in every patient but on average in one out of every two patients.
3. As a last-resort method, LD-RT is clearly superior to all other local conservative therapies in terms of both scope and duration of effect.
4. By involving nearly all practicing colleagues in research, they can be promptly informed of the overall results. This is another benefit of the DROM method.

## Data Availability

www.arthose.de

https://www.arthrose.de

